# The association of blood-based biomarkers of neuropathology with cognitive performance and incident dementia in a diverse, nationally-representative sample of US adults

**DOI:** 10.64898/2026.01.07.26343604

**Authors:** Jessica D. Faul, Eileen M. Crimmins, Jung Ki Kim, Bharat Thyagarajan, Jonathan W. King, David R. Weir, Kenneth M. Langa

## Abstract

**INTRODUCTION:** The association of blood-based biomarkers of neuropathology with cognition, dementia, and mortality and how these association potentially differ by race/ethnicity, has not been examined in large, diverse, nationally-representative samples of adults.

**METHODS:** The sample included Health and Retirement Study (HRS) respondents over age 50 with blood-based neuropathology biomarker, demographic, and cognitive data (n=4,214). A**β**-40, A**β**-42, neurofilament light chain (NfL), and glial fibrillary acidic protein (GFAP) were measured in plasma (Quanterix Neurology 4-Plex E kit), and phosphorylated tau (pTau-181) was measured in serum (Quanterix Advantage V2.0 kit). Cognitive tests included immediate and delayed word recall, serial 7s, and backward counting (total score 0-27). Dementia classification relied on a diagnostic algorithm previously validated in the HRS.

**RESULTS:** When each biomarker was analyzed individually, higher A**β**-42/A**β**-40 ratio was associated with better cognitive function among non-Hispanic (NH) whites. Higher NfL was associated with worse cognitive function in the total sample and in each race/ethnic group (NH white, NH black, and Hispanic). Higher pTau-181 was associated with worse cognitive function in the total and NH white sample. Higher GFAP was related to worse cognitive function in the total sample only. In a model that included all four biomarkers, NfL remained significantly related to cognitive performance in the total sample and in each race/ethnic group, and irrespective of APOE status. NfL was predictive of 6-year incident dementia in our sample (OR=1.33). All four markers significantly predicted 6-year mortality.

**DISCUSSION:** In a large nationally-representative sample of US adults, we found that NfL was the most consistent predictor of cross-sectional and incident dementia 6 years post blood collection. NfL was also the most consistent predictor across race/ethnic groups examined in our study.

**Highlights:** There are currently limited data on blood-based biomarkers of neuropathology as predictors of cognitive performance and incident dementia in diverse, population-based cohort studies.

We used data from the Health and Retirement Study (n-=4,214) to examine the association between blood-based biomarkers of neuropathology and cognitive function, as well as their association with incident dementia and mortality 4 years after measurement.

Mean levels of A**β**-42/A**β**-40 were similar across race/ethnic groups and age groups in this US population-representative sample where selection effects have been minimized. Average NfL was higher among non-Hispanic blacks and Hispanics; GFAP was higher among non-Hispanic blacks as compared to non-Hispanic whites.

In a model that included all four biomarkers, NfL remained significantly related to cognitive performance in the total sample and in each race/ethnic group.

NfL was associated with incident dementia 6 years after measurement in the total sample. A**β**-42/A**β**-40 ratio was predictive of 6-year incident dementia among those with at least one APOE e4 allele.

## 1. Background

Ongoing enhancements in imaging, especially PET and cerebrospinal fluid (CSF) diagnostics, have allowed for a better understanding of the neuropathologies associated with the onset and progression of Alzheimer’s disease and related dementias (ADRD). However, these diagnostic methods are often costly and are more invasive than a traditional blood draw, and consent rates (especially for the lumbar puncture required to collect CSF) can be quite low even in volunteer cohorts [1]. Hence, they remain difficult to implement in large-scale, epidemiological studies or for widespread population screening for ADRD. Thus, the development of reliable and valid blood-based biomarkers for ADRD is an area of growing interest for many researchers and essential to the advancement of ADRD diagnosis and treatment [2–7].

Blood-based biomarkers associated with neuropathology are notably difficult to detect due to low levels in the blood. This is likely related to the function of the blood-brain barrier, which tightly regulates movement of ions and molecules including proteins in and out of the blood vessels that vascularize the central nervous system. This regulation is essential for adequate neuronal function, but it leads to significantly lower concentrations of brain-related proteins in the blood [8, 9]. Thus, assays with high analytical sensitivity are needed for reliable detection of blood-based biomarkers associated with cognitive impairment [10].

Among potential blood-based biomarkers for ADRD, Amyloid-beta 40 (Aβ-40), Amyloid-beta 42 (Aβ-42), neurofilament light chain (NfL), Glial Fibrillary Acidic Protein (GFAP), and the species of tau phosphorylated at threonine 181 (pTau-181) have been used to detect early-stage disease in various clinical samples [8–12]. Amyloid-beta 40 and Amyloid-beta 42 are peptides that accumulate in the brain and are implicated in the early pathogenesis of neurodegeneration. The ratio of these two proteins, Aβ42/40 has been longitudinally associated with greater decline in cognitive function and amyloid deposition in the brain. Other research has demonstrated the utility of using plasma Aβ42/40 to detect early-stage Alzheimer’s disease [13]. Plasma Aβ42/Aβ40 has also been shown to be associated with progression to both MCI and AD dementia from baseline [14, 15]. Neurofilament light (NfL) is also associated with neurodegenerative disease, due to its role in axonal damage. Glial Fibrillary Acidic Protein (GFAP) is associated with mild cognitive impairment and AD. Tau proteins have been shown to increase as dementia progresses and increased pTau-181 has been seen in neurofibrillary tangles associated with AD [16].

The establishment of a biomarker-based diagnostic classification for Alzheimer’s disease (the Amyloid, Tau, Neurodegeneration, or ATN framework) [17] and the recent FDA approval of monoclonal antibodies (lecanemab, marketed as Leqembi, and donanemab, marketed as Kisunla) aimed at reducing amyloid build-up in the brain, have created a pressing need to better understand the prevalence and prognosis of different ATN biomarker profiles in more representative, population-based samples, especially those with sufficient numbers of participants from minoritized populations. Few population-based cohort studies have investigated the association between blood-based biomarkers and cognitive outcomes to determine whether they could be used to identify individuals at risk for cognitive decline without requiring the costly collection of brain and cerebrospinal fluid biomarkers [18, 19]. Moreover, given previous work suggesting that minoritized groups in the US may be at greater risk of ADRD, it is important to test associations discovered in predominantly white populations in non-white populations; however, most previous research in this area has used samples with very limited representation of non-white population groups [20]. Recent work has examined the distribution of AD biomarker concentrations in a US based cohort of middle-aged adults but did not examine the association with cognitive function, dementia incidence, or the distribution of these markers across a wider age group of older adults [21].

## 2. Methods

### 2.1 Participants

Data come from the 2016 Health and Retirement Study (HRS) Venous Blood Study (VBS). The HRS is a nationally-representative panel study of Americans aged 51 and older with extensive survey data, including cognitive functioning measures, collected every two years. The HRS includes oversamples of black and Hispanic persons [22–25]. All respondents who were in the panel and completed a self-interview in 2016 were invited to participate in the VBS (n = 9,934). The sample for this study was selected from 2016 HRS respondents born 1959 and earlier who also participated in the 2016 VBS. From this group, a probability sample was selected from among respondents who were 65 and older and thus eligible for the 2016 Harmonized Cognitive Assessment Protocol (HCAP) or who are eligible for a future HCAP (a random half of HRS sample members not yet 65 years old). Neuropathological markers were assayed on this representative subset of HRS participants (n=4,214).

### 2.2. Blood Collection

Blood was collected from consenting participants in their home by a certified phlebotomist. Samples were shipped overnight in temperature-controlled packages to the receiving lab, the Advanced Research and Diagnostics Laboratory (ARDL), at the University of Minnesota [26]. Prior pilot work performed by this lab established the stability and validity of measuring blood-based biomarkers of neuropathology using this collection protocol [27]. In this collection wave, 92% of the samples arrived in the lab within 24 hours of collection and 99% within 48 hours.

### 2.3 Measures

#### 2.3.1 Blood-based biomarkers of neuropathology

A**β**-40, A**β**-42, NfL, and GFAP were measured in plasma using the Quanterix Neurology 4-Plex E kit. Phosphorylated tau (pTau-181) was measured in serum using the Quanterix Advantage V2.0 kit. All measurements were conducted on the same HD-X analyzer (Quanterix Inc.). Due to its digital quantification of protein concentrations, the Quanterix Simoa immunoassay is approximately 1000-fold more sensitive than conventional ELISA approaches [28, 29]. The assays were run in three batches using the same reagent lot. Based on pooled plasma and serum controls analyzed across the analytical runs, the coefficient of variation ranged from 8.55% to 13.34%.

#### 2.3.2 APOE e4

APOE genotyping was performed on salivary DNA samples using TaqMan allelic discrimination SNP assays, C_ 3084793 (rs429358) and C_ 904973 (rs7412) using a 7900HT Sequence Detection System and quantified using SDS v2.1software (Applied Biosystems, Foster City, CA). Genotypes are determined using TaqMan Genotyper software. For participants for whom direct genotyping was not available, either because there was insufficient sample or the sample did not pass QC (where one or both of the SNPs that determine the APOE isoform failed), APOE status was imputed from previously genotyped array data using the Illumina HumanOmni2.5 array. Imputations referenced the 1000 Genomes Cosmopolitan Reference Panel (phase 3). Dosage data for rs7412 and rs429358 was used to assign the “best guess genotypes” for each respondent in in turn to infer the APOE isoform. The DNA samples were genotyped and analyzed at the Center for Inherited Disease Research (CIDR) Genetic Resources Core Facility (GRCF) and Fragment Analysis Facility (FAF) at Johns Hopkins University (https://cidr.jhmi.edu). These APOE data are available from the HRS [30].

### 2.3.3 Cognitive function

Cognitive function was assessed in 2016 at the time of blood collection using selected items from the modified version of the Telephone Interview for Cognitive Status (TICS-m), including immediate and delayed recall of 10 nouns, serial 7s subtraction, and a backward count. These items sum to a range of 0-27 points. This battery is a validated instrument used to screen for dementia and mild cognitive impairment [31, 32]. We used the imputed cognition data released by HRS in order to minimize the effects of item-level non-response among respondents [33]. Cognitive function was also assessed in those who participated in the 2018, 2020, and 2022 waves of HRS (2022 being the most current wave for which imputed data are available).

### 2.3.4 Dementia status and incident dementia

Dementia status was defined using an algorithmic-based approach that has previously been validated in this sample [32]. This algorithm, the Langa-Weir method, uses the TICS-m score or information from a proxy report, usually from a spouse/partner or family member), about difficulty with instrumental activities of daily living (IADLs) and assessment of respondent’s memory as well as the interviewer’s assessment of the respondent’s cognition to create a total cognition score based on proxy information. Because cognitive decline is a common cause of study attrition, the use of proxy reports reduces the bias that would result from excluding those participants [34]. Diagnoses derived using this algorithm have been used extensively in the literature focusing on population-level effects in lieu of a clinically-determined dementia diagnosis [32]. More information on this measure is available from the HRS website [35]. Incident dementia was determined as a transition from normal cognitive functioning or cognitive impairment, not dementia (CIND) to dementia status at any time in the 6-year observation period after 2016. Only respondents who were never classified as having dementia in 2016 were included in the analysis of incidence (n=3,915). To adjust for mode effects and slightly higher performance among respondents who answered the survey on the web, the criteria for CIND classification was adjusted by one point [36].

Proxy interviewers are used in the HRS if the respondent is unable or unwilling to complete the survey on their own. Proxy respondents are asked to rate the respondent’s overall memory, change in memory compared to the prior wave, and their behavior in terms of overall judgment and organization of daily life. These questions are adapted from the short form of the Informant Questionnaire on Cognitive Decline in the Elderly [37, 38](IQCODE). While the cognitive measures used for proxy and self-respondents are not the same, each can each be used to assign the same broad categories of cognitive impairment and dementia [32].

#### 2.2.5 Mortality

HRS monitors vital status through efforts to locate respondents and through administrative linkages to the National Death Index. Mortality coverage is essentially complete [39]. Six-year mortality was determined using vital status known to HRS as of the 2018, 2020, and 2022 survey waves.

#### 2.2.6 Covariates

Age, sex/gender, race/ethnicity (non-Hispanic black, Hispanic, and non-Hispanic white) were ascertained from the survey.

### 2.3 Analyses

We examined the mean and range of each neuropathological biomarker in total and by race/ethnic group. Next we performed ordinary least squares regressions of cross-sectional cognitive function or dementia status on each neuropathological marker adjusting for age, sex, and batch. We analyzed continuous outcome data using OLS regression. We analyzed binary outcome data, 6-year incident dementia, and mortality, using logistic regression among respondents without dementia in 2016 (n=3,911) using logged and standardized versions of the blood-based markers. Models were also run separately by race/ethnic group. Additional models also adjusted for APOE e4 allele count and were run separately by APOE e4 status (no e4 alleles vs. any e4 alleles). To determine whether kidney function affected our results, we also ran a parallel set of models adjusting for estimated glomerular filtration rate (eGFR) estimated using sex-specific models using serum creatinine with age and race adjustment.

All models were run using SAS 9.4.

## 3. Results

### 3.1 Descriptive Statistics

The majority of the sample was female (54.3%) with an average age of 68.1 years (Supplementary Table 1). The sample was largely non-Hispanic white (n=2,850, 80.2%); however, it included sizable samples of non-Hispanic black (n=726, 10.3%) and Hispanic participants (n=638, 9.5%). The average cognitive score in 2016 was 15.67. In 2016, 83.7% of the sample was classified as cognitively normal with 13.2% classified as having CIND and 3.1% with dementia. In the sample with measured APOE e4 status (n=3,755), 75.8% had no APOE e4 risk alleles while 22% had one e4 allele and 2.3% had two e4 alleles. By 2022, 3.6% of the sample experienced dementia onset. The overall 6-year mortality rate was 13.7%.

The average A**β**-42/A**β**-40 ratio was 0.066 (SD=0.024) with a minimum of 0.012 and a maximum of 0.584 (Supplementary Table 1). The mean A**β**-42/A**β**-40 ratio did not differ across race/ethnic groups, but the SD for the non-Hispanic white group was much higher than among the non-Hispanic black and Hispanic groups (Table 1). Mean NfL in the total sample was 23.3 (SD=25.94), mean pTau-181 was 2.01 (SD=2.11) and GFAP was 99.49 (SD=72.08) (Supplementary Table 1). Compared to non-Hispanic whites, non-Hispanic Blacks had higher NfL (25.6 vs 22.6 for NH white, p=0.0183) and GFAP (109.2 vs. 98.7 for NH white, p=0.0062); and Hispanics had higher NfL (26.4 vs. 22.6 for NH white, p=0.0032)(Table 1 and Supplementary Figure 1a-d). NfL, pTau-181, and GFAP all increased with age while no age gradient was observed for A**β**-42/A**β**-40 (Supplementary Table 2). A**β**-42/A**β**-40 was not correlated with NfL or GFAP but was somewhat negatively correlated with pTau-181. NfL was moderately positively correlated with pTau-181 and GFAP. GFAP was also moderately positively correlated with pTau-181 (Table 2).

### 3.2 Cross-sectional results

Table 3 shows the individual and combined linear regressions of the neuropathological markers on cognitive function measured cross-sectionally. In the total sample, all four markers were significantly associated with cognition with the A**β**-42/A**β**-40 ratio having a positive relationship with cognition while NfL, pTau-181, and GFAP were inversely related to the cognitive functioning score. In the non-Hispanic white sample all makers with the exception of GFAP were associated with the cross-sectional cognitive test score. In the samples of non-Hispanic blacks and Hispanics, only NfL was significantly associated with cognitive function. In the combined model, in which all four markers were considered at once, only NfL remained significantly associated with cognitive performance in the total sample and across all three race/ethnic groups.

In the models that predict dementia status cross-sectionally, NfL and GFAP were associated with a higher odds of dementia in the total sample. In the combined models, only NfL remained associated with a higher odds of dementia (OR=1.26 [95% CI 1.00, 1.59)]) and only in the total sample (Table 4).

### 3.3 Considering APOE e4

The number of APOE e4 of alleles was negatively associated with cognitive performance in all race/ethnic groups, but only significantly so in the total, NH white, and Hispanic samples (Supplementary Table 3). In the combined models, having any APOE e4 alleles was associated with cognitive performance in the total, NH white, and Hispanic samples. NfL remained predictive of cognition across groups with APOE e4 allele count in the model while GFAP was significantly associated with cognitive performance only in the total and NH white samples.

Supplementary Table 4 shows the results stratified by APOE status, in those with and without any APOE e4 alleles. NfL was associated with cognitive function irrespective of APOE status. pTau-181 was associated with cognitive performance in those with any APOE e4 alleles. In the combined model in which the blood-based markers were considered simultaneously NfL and GFAP were associated with cognitive performance irrespective of APOE status.

### 3.4 Longitudinal results

Table 5 and Figure 1 show the results of the multinomial logistic regression predicting incident dementia and death 6 years after the neuropathological biomarkers were assessed. Higher A**β**-42/A**β**-40 ratio and GFAP were associated with a reduced odds of death (0.81 and 0.66, respectively), while p-Tau-181 and NfL were both positively associated with mortality risk (OR=1.15 and 2.32, respectively). Higher NfL was predictive of 6-year mortality and in those with and without APOE e4 alleles while a higher A**β**-42/A**β**-40 ratio and higher GFAP were protective of mortality in those with and without APOE e4 alleles. NfL was predictive of 6-year incident dementia in our sample (OR= 1.36). In the sample stratified by APOE e4, only the A**β**-42/A**β**-40 ratio predicted incident dementia 6 years later (OR=0.68) and only among those with at least one APOE e4 allele.

### 3.5 Kidney function

Adjusting for eGFR using a serum creatinine-based formula did not change the results described above. These results are shown in Supplementary Table 5.

## 1. Discussion

In a large nationally-representative sample of US adults, we found that NfL was the most consistent predictor of cross-sectional and incident dementia 6 years post blood collection. NfL was also the most consistent predictor across race/ethnic groups examined in our study. There has been a substantial interest in examining how biomarkers can be used to aid in early detection and better diagnosis of ADRD. The development and recent FDA approval of monoclonal antibodies aimed at removing cerebral amyloid and delaying the progression of AD has heightened the need for easier and earlier identification of biomarker profiles associated with increased risk of cognitive decline and dementia. Indeed, population screening to identify at-risk older adults for further evaluation and potential treatment is a goal that we’d like to achieve in the near term [40]. However, to realize this goal the biomarkers must be not only accurate, but easy and cost-effective to collect. There are numerous studies that examine the association between CSF/neuroimaging biomarkers and neuropathology and even more that show that blood-based biomarkers are strongly associated with CSF/neuroimaging biomarkers, but there are very few that examine feasibility and utility of assessing blood-based biomarkers in any population-based sample, let alone a diverse one that is reflective of the US population [41, 42]. This study represents the first time that such extensive biomarker data have been collected in a large nationally-representative, population-based survey with both cross-sectional and longitudinal cognitive assessment.

In our sample, correlations were modest among pTau-181, NfL, and GFAP. Aβ42/40 was weakly and negative correlated with pTau-181. These relationships confirm that the markers examined in this study are likely measuring different pathological pathways in the brain Mean levels of Aβ42/40 were similar across race/ethnic groups. This contrasts with prior research that has shown different levels of Aβ42 and Aβ40 among NH black study participants [21, 43]. In our sample, non-Hispanic Blacks had higher NfL and GFAP compared to non-Hispanic whites, indicating higher brain pathology. Hispanics also had higher NfL compared to non-Hispanic whites. In another representative sample of US-based adults ages 56-63 years using the same analytic platforms, the patterns were quite different with blacks having a lower mean Aβ42/40 ratio and lower mean NfL than whites and Hispanics having lower GFAP on average as compared to whites. Our patterns held even when restricting our sample to respondents ages 56-63.

Aβ42/Aβ40 ratio was associated with cognitive performance in the NH white sample only, in agreement with previous research [12, 18, 44]. NfL, pTau-181 and GFAP were all individually associated with cross-sectional cognitive performance in the total sample. NfL was the only neuropathological marker that was associated with cognitive performance across all race/ethnic groups. In a model that included all four biomarkers simultaneously, NfL remained significantly related to cognitive performance in the total sample and across all race/ethnic groups. NfL is a more generalized biomarkers of neuronal injury and inflammation so may be better at reflecting cognitive decline in the population resulting from multiple etiologies. pTau-181 was predictive of cognitive performance on its own but not when considered with the other neuropathological markers. More recent papers have examined pTau-217 and pTau-231 as potentially diagnostically superior to p-Tau 181 [45] and more indicative of amyloid plaques in the brain [46]. If more evidence confirms this to be true, future research could consider these analytes now that commercially available kits are becoming more readily available.

In contrast to other work, our findings do not indicate that plasma Aβ42/40 has a high screening value in identifying incident dementia onset, except in those who also have at least one APOE e4 allele. This would fit the idea that prior work using clinic-based non-representative samples suffer from a “spectrum bias” and in those selected samples the biomarkers are likely to have better predictive power than in a nationally-representative sample with a wider range of neuropathology and comorbidities. NfL was the strongest predictor of incident dementia 6 years later, although the relationship was harder to discern once the sample was stratified by APOE risk alleles. All of the markers were predictive of mortality in the total sample and surprising the relationship between GFAP and mortality was in the unexpected direction with higher GFAP associated with lower odds of death in 6 years.

This study was limited by the lack of PET or CSF biomarker level measurement as methods of confirmation or additional diagnostic specificity. In HRS, dementia classification and incidence is determined from an algorithm based on cognitive test performance. The algorithm was developed using an expert panel with access to medical records, and while used widely, results may differ from a physician diagnosis. Generally, the lack of diagnostic specificity is a challenge to the development of blood-based assessment or screening protocols for ADRD. Cognitive decline and dementia are typically due to multiple and overlapping neuropathologies [47] which makes both accurate diagnosis and prediction of prognosis more difficult. However, this mirrors the difficulty that will exist when blood-based tests are used to screen the general population. More research will be needed to determine exactly which markers or combination of markers map to each disease pathology.

A strength of this study is that we used immunoassay methods from Quanterix, which are widely available and relatively inexpensive. This platform is likely more similar to the technology that hospitals and clinics may have access to for assessment as compared to the immunoprecipitation-mass spectrometry (IP-MS) methods used for Aβ42 and Aβ40 measurement that are only accessible in select research and clinical laboratories. There are several platforms available and in development for the assessment of AD/ADRD-related markers. These include the traditional Elisa techniques, the aforementioned Quanterix and IP-MS, as well as multiplex immunoassay kits available from Meso Scale Discovery (S-Plex and a new ultrasensitive p-Tau 217 kit), the Alamar NULISA platform, and the Roche Elecsys ® kits using the Cobas analyzer [48]. Future research should include a cross-platform analysis so that results across studies can be interpreted more easily relative to one another.

Due to the constraints of our blood collection protocol, specifically that the plasma was unable to be processed until it arrived at the central laboratory 24-48 hours after collection, it was not possible to examine p-Tau using plasma or to examine Aβ42 and Aβ40 plasma levels as individual markers. In our pilot work we found that a delay in processing and exposure to +18°C affected Aβ40 and Aβ42 concentrations while the Aβ42/Aβ40 ratio was unaffected[27]. Blood collection in a geographically dispersed sample is challenging and some compromises have to be made to make it feasible. However, our earlier work as well as this study show that assessment using field-collected blood samples, even with 24-48 hour delayed processing, is possible and valid using serum for p-Tau assessment and plasma for the measurement of NfL, GFAP, and the Aβ42/Aβ40 ratio. Hopefully, this means that more population-based and studies with greater racial and ethnic diversity will be able to contribute to this research without the constraints of clinic-like blood collection protocols.

The main strength of this study is the use of a large, nationally-representative, and diverse sample with longitudinal assessment. Prior population-based samples have been used in biomarker assessment research but were comprised of volunteers or selected from voter registration lists and consist largely of non-Hispanic white participants [18]. This project adds assessment of these markers in a diverse sample including non-Hispanic black and Hispanic participants.

In summary, we have shown that neuropathological biomarkers can be measured in population studies with appropriate protocols and that these markers relate to cognitive outcomes, both cross-sectionally and longitudinally. In our study, NfL was the strongest predictor of cognitive performance and concurrent dementia across race/ethnic groups and irrespective of APOE e4 allele status and kidney function, as well as of dementia 6 years after assessment. These results show the importance of examining blood-based biomarkers of neuropathology as predictors of cognitive performance and dementia in diverse, population-based samples. Examination of these relationships in more studies that include large samples of non-Hispanic black and Hispanic is warranted before making recommendations for wide-spread biomarker-based screening in clinical practice. In addition, examining longitudinal change in these markers will be useful indicators of sub-clinical disease progression.

## Data Availability

All data referenced are available online at https://hrsdata.isr.umich.edu/

## 6. Acknowledgements/Conflicts/Funding Sources

The HRS (Health and Retirement Study) is sponsored by the National Institute on Aging (grant number NIA U01AG009740) and is conducted by the University of Michigan. Research reported in this study was also supported by NIA U01 AG058499-06 and The Alzheimer’s Association HRS-18-586069C.

One co-author of this paper (JWK) is an employee of the National Institute on Aging (NIA), but the opinions expressed are their own and not those of the NIA, the National Institutes of Health, or the U.S. Department of Health and Human Services.

