## Supplementary material for "The association of blood-based biomarkers of neuropathology with cognitive performance and incident dementia in a diverse, nationally-representative sample of US adults": Tables and Figures

**Table 1: Age and Sex Adjusted Means (Unadjusted SD, Min, Max) and Distributions of Neuropathological Biomarkers by Race/Ethnic Group**

| | Non-Hispanic White (n=2,850) | | | | Non-Hispanic Black (n=726) | | | | Hispanic (n=638) | | | |
|---|---|---|---|---|---|---|---|---|---|---|---|---|
| | **Mean** | **SD** | **Min** | **Max** | **Mean** | **SD** | **Min** | **Max** | **Mean** | **SD** | **Min** | **Max** |
| **AB42/40 Ratio** | 0.066 | 0.027 | 0.012 | 0.584 | 0.068 | 0.013 | 0.017 | 0.351 | 0.068 | 0.015 | 0.017 | 0.200 |
| **NfL** | 22.64 | 21.81 | 3.0 | 373.68 | 27.35 | 31.85 | 1.7 | 635.3 | 28.24 | 34.03 | 3.5 | 897.1 |
| **pTau-181** | 2.05 | 2.25 | 0.028 | 43.00 | 2.08 | 1.67 | 0.028 | 40.08 | 2.02 | 1.90 | 0.028 | 32.89 |
| **GFAP** | 98.0 | 76.80 | 8.9 | 789.1 | 114.14 | 68.14 | 14.9 | 682.9 | 101.24 | 51.13 | 5.7 | 414.25 |

Age and sex adjusted

**Table 2: Correlations Among Neuropathological Biomarkers (N=4,214)**

| | AB42/40 Ratio | NfL | pTau-181 | GFAP |
|---|---|---|---|---|
| **AB42/40 Ratio** | | -0.0027 | -0.0378* | 0.0007 |
| **NfL** | | | 0.2962*** | 0.4644*** |
| **pTau-181** | | | | 0.2989*** |
| **GFAP** | | | | |

***p<.001; **p<.01; *p<.05

**Table 3: Linear Regressions of Each Neuropathological Biomarker on Cross-sectional Cognitive Functioning Score (n=4,214)**

| | Total | | Non-Hispanic White | | Non-Hispanic Black | | Hispanic | |
|---|---|---|---|---|---|---|---|---|
| **Cognitive functioning score (0-27)** | | | | | | | | |
| | **b** | **p** | **b** | **p** | **b** | **p** | **B** | **p** |
| **zAB42/40** | 0.15 | 0.0192 | 0.20 | 0.0064 | -0.10 | 0.5823 | -0.09 | 0.6267 |
| **zNfL** | -0.63 | <.0001 | -0.63 | <.0001 | -0.65 | <.0001 | -0.57 | 0.0013 |
| **zpTau-181** | -0.21 | 0.0023 | -0.28 | 0.0005 | -0.07 | 0.6665 | -0.27 | 0.1168 |
| **zGFAP** | -0.17 | 0.0444 | -0.12 | 0.1920 | -0.11 | 0.5817 | -0.22 | 0.3203 |
| **Combined model** | | | | | | | | |
| **zAB42/40** | 0.13 | 0.0416 | 0.18 | 0.0142 | -0.11 | 0.5212 | -0.15 | 0.4423 |
| **zNfL** | -0.67 | <.0001 | -0.65 | <.0001 | -0.87 | <.0001 | -0.58 | 0.0048 |
| **zpTau-181** | -0.06 | 0.4357 | -0.16 | 0.0621 | 0.21 | 0.2432 | -0.12 | 0.5220 |
| **zGFAP** | 0.15 | 0.0918 | 0.19 | 0.0720 | 0.38 | 0.0911 | 0.12 | 0.6314 |

Models adjusted for age, sex, and batch

Neuropathological biomarkers are logged and standardized to facilitate comparison.

| Table 4: Logistic Regressions of Each Neuropathological Biomarker on Cross-sectional Predicted Dementia Status (n=4,214) | | | | | | | | | | | | |
|---|---|---|---|---|---|---|---|---|---|---|---|---|
| | Total | | | Non-Hispanic White | | | Non-Hispanic Black | | | Hispanic | | |
| **Predicted dementia** | | | | | | | | | | | | |
| | **OR** | **95% CI** | **p** | **OR** | **95% CI** | **p** | **OR** | **95% CI** | **p** | **OR** | **95% CI** | **p** |
| **zAB42/40** | 0.92 | 0.76, 1.12 | 0.4016 | 0.77 | 0.59, 1.00 | 0.0478 | 0.88 | 0.57, 1.37 | 0.5822 | 1.11 | 0.76, 1.62 | 0.5892 |
| **zNfL** | 1.34 | 1.10, 1.64 | 0.0040 | 1.48 | 1.10, 1.99 | 0.0093 | 1.40 | 1.01, 1.94 | 0.0434 | 0.84 | 0.52, 1.36 | 0.4754 |
| **zpTau-181** | 1.13 | 0.92, 1.38 | 0.2473 | 1.45 | 1.11, 1.90 | 0.0066 | 1.08 | 0.70, 1.67 | 0.7307 | 0.81 | 0.52, 1.26 | 0.3488 |
| **zGFAP** | 1.32 | 1.04, 1.69 | 0.0234 | 1.47 | 1.05, 2.06 | 0.0255 | 1.30 | 0.80, 2.11 | 0.2904 | 0.96 | 0.55, 1.69 | 0.8851 |
| **Combined model** | | | | | | | | | | | | |
| **zAB42/40** | 0.94 | 0.78, 1.14 | 0.5530 | 0.81 | 0.61, 1.06 | 0.1172 | 0.90 | 0.58, 1.41 | 0.6507 | 1.07 | 0.72, 1.58 | 0.7470 |
| **zNfL** | 1.26 | 1.00, 1.59 | 0.0494 | 1.26 | 0.89, 1.79 | 0.1917 | 1.41 | 0.96, 2.08 | 0.0820 | 0.85 | 0.49, 1.47 | 0.5635 |
| **zpTau-181** | 1.01 | 0.82, 1.26 | 0.9143 | 1.30 | 0.97, 1.75 | 0.0763 | 0.89 | 0.56, 1.42 | 0.6179 | 0.84 | 0.52, 1.37 | 0.4797 |
| **zGFAP** | 1.16 | 0.88, 1.53 | 0.2946 | 1.15 | 0.78, 1.69 | 0.4743 | 1.07 | 0.62, 1.86 | 0.8026 | 1.10 | 0.58, 2.10 | 0.7740 |

Models adjusted for age, sex, and batch

Neuropathological biomarkers are logged and standardized to facilitate comparison.

| Table 5: Multinomial Logistic Regressions of 6-year Incident Dementia and Death by APOE e4 Status | | | | | | | | | | | | |
|---|---|---|---|---|---|---|---|---|---|---|---|---|
| | | | | | Without APOE e4 | | | | With APOE e4 | | | |
| | Dementia Onset in 6 years | | Death in 6 years | | Dementia Onset in 6 years | | Death in 6 years | | Dementia Onset in 6 years | | Death in 6 years | |
| | (n=3,915) | | (n=3,915) | | (n=2,625) | | | | (n=864) | | | |
| | **OR** | **95% CI** | **OR** | **95% CI** | **OR** | **95% CI** | **OR** | **95% CI** | **OR** | **95% CI** | **OR** | **95% CI** |
| **zAB42/40** | 1.01 | 0.85-1.22 | 0.81*** | 0.72-0.90 | 1.11 | 0.87-1.41 | 0.79** | 0.69-0.91 | 0.68* | 0.48-0.96 | 0.76* | 0.59-0.97 |
| **zNfL** | 1.36* | 1.05-1.75 | 2.32**** | 2.00-2.68 | 1.09 | 0.76-1.58 | 2.18**** | 1.84-2.58 | 1.25 | 0.75-2.10 | 2.53**** | 1.79-3.58 |
| **zpTau181** | 1.14 | 0.93-1.41 | 1.15* | 1.02-1.30 | 1.13 | 0.86-1.50 | 1.11 | 0.96-1.28 | 1.36 | 0.88-2.10 | 1.14 | 0.82-1.58 |
| **zGFAP** | 1.29 | 1.00-1.67 | 0.66**** | 0.57-0.77 | 1.14 | 0.79-1.64 | 0.66**** | 0.55-0.79 | 1.01 | 0.59-1.72 | 0.61** | 0.42-0.88 |

Models adjusted for age, sex, and batch
Neuropathological values are logged and standardized to facilitate comparison
****p<.0001; ***p<.001; **p<.01; *p<.05

**Figure 1. Multinomial Logistic Regressions of 6-year Incident Dementia and Death by APOE e4 Status**

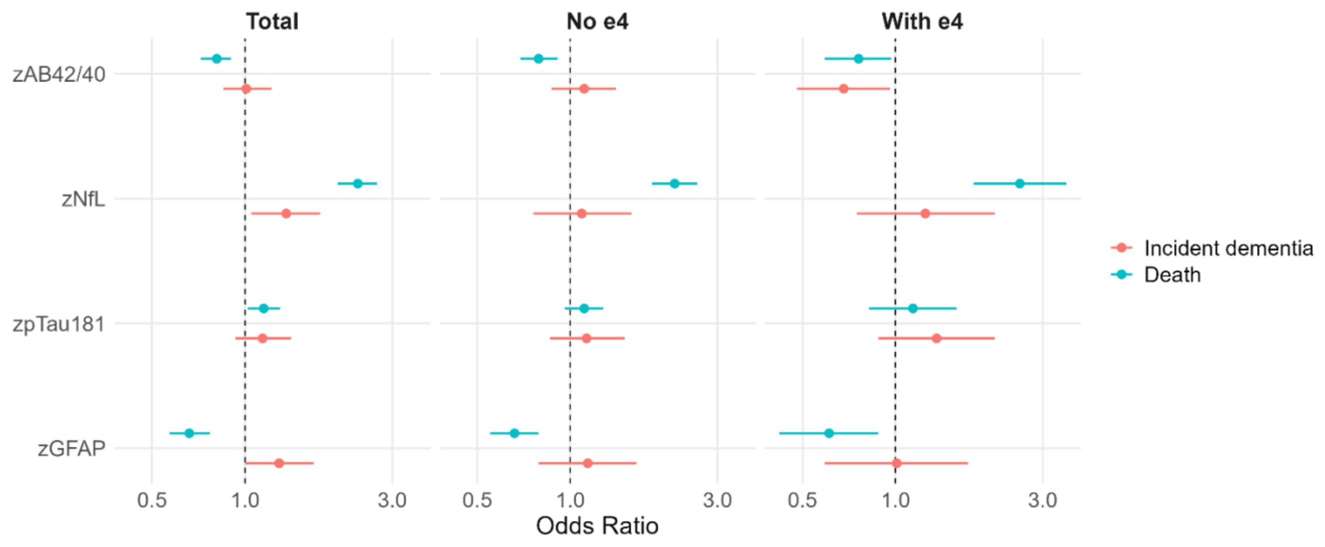

Models adjusted for age, sex, and batch
Neuropathological values are logged and standardized to facilitate comparison

**Supplementary Table 1: Descriptive Statistics of the 2016 HRS Sample with Neuropathological Biomarkers (N=4,214)**

| | Mean / % | SD | Min | Max |
|---|---|---|---|---|
| **AB42/40 Ratio** | 0.066 | 0.024 | 0.012 | 0.584 |
| **NfL** | 23.30 | 25.94 | 1.7 | 897.1 |
| **pTau-181** | 2.01 | 2.11 | 0.028 | 43.00 |
| **GFAP** | 99.49 | 72.08 | 5.7 | 789.1 |
| **Age (yrs)** | 68.1 | 9.35 | 50 | 100 |
| **Female** | 54.3% | | | |
| **Race/ethnicity** | | | | |
| **NH White (n=2850)** | 80.2% | | | |
| **NH Black (n=726)** | 10.3% | | | |
| **Hispanic (n=638)** | 9.5% | | | |
| **Education (hrs)** | | | | |
| **0-12 yrs** | 43.7% | | | |
| **13-15 yrs** | 25.9% | | | |
| **16+ yrs** | 30.4% | | | |
| **Batch** | | | | |
| **Batch 1** | 21.3% | | | |
| **Batch 2** | 30.3% | | | |
| **Batch 3** | 48.4% | | | |
| **Cognitive Status 2016** | | | | |
| **Normal** | 83.7% | | | |
| **CIND** | 13.2% | | | |
| **Dementia** | 3.1% | | | |
| **Cognitive Score 2016** | 15.67 | 4.33 | 0 | 27 |
| **Incident Dementia in 6 years (n=3,915)** | | | | |
| **No Onset** | 82.7% | | | |
| **Onset of Dementia (n=196)** | 3.6% | | | |
| **Death (n=602)** | 13.7% | | | |
| **APOE e4 (n=3,755)** | | | | |
| **0** | 75.8% | | | |
| **1** | 22.0% | | | |
| **2** | 2.3% | | | |
